# Blood pressure self-monitoring practice and associated factors among adult hypertensive patients on follow-up at South Wollo Zone Public Hospitals, Northeast Ethiopia

**DOI:** 10.1101/2023.02.02.23285364

**Authors:** Esmael Geleta, Zemen Mengesha, Belachew Tegegne, Sewunet Ademe, Tiliksew Liknaw, Afework Edmealem

## Abstract

**Background:** Hypertension is a silent killer that causes serious health issues in all parts of the world.It is risk factor for cardiovascular disease, stroke, and kidney disease. Self-monitoring practice has been identified as an important component of hypertension management. Hence, this study aimed to assess blood pressure self-monitoring practice and associated factors among adult hypertensive patients on follow-up in South Wollo Zone public hospitals, Northeast Ethiopia.

**Methods:** Hospital based cross-sectional study was conducted from June 1 to June 30, 2022, among 336 adult hypertensive patients on follow-up at selected South Wollo Zone public hospitals.Data were collected by using self-administered pre-tested structured questionnaires; the collecteddata were entered into Epi-data version 4.6 and then exported to SPSS version 25 software for analysis. Descriptive statistics such as frequency and percentage were used to describe the study participants. Tables and texts were used for data presentation. Binary logistic regression was conductedto test the association between the independent and dependent variables. Adjusted Odds Ratio with 95% CI was estimated to identify the factors associated with BP self-monitoring and thelevel of significance wasdeclared at P-value <0.05.

**Results:** The proportion (95% CI) of Blood Pressure Self-Monitoring Practice among hypertensive patients in South Wollo Zone Public Hospitals was 8.93% [95%CI; 6.3, 12.5]. In the multivariable analysis, urban residence [AOR=3.97, 95%CI (1.11, 14.20)], comorbidity [AOR=4.80, 95%CI (1.23, 18.69)], regular healthcare professional visit [AOR=4.64, 95%CI (1.02, 21.14)], advice on the type of devices used for BP self-monitoring [AOR=5.26, 95%CI (1.49, 18.58)], and knowledge on hypertension self-care [AOR=13.13, 95%CI (4.21, 40.99)] were positively associated with BP self-monitoring practice.

**Conclusion:** In the study, the proportion of Blood Pressure Self-Monitoring Practice was low.Living in urban areas, comorbidity, regular healthcare professional visits, advice on the type of devices used for Blood Pressure self-monitoring, and knowledge of hypertension self-care were positively associated with Blood Pressure Self-Monitoring Practice.

## Introduction

Hypertension is a condition in which the blood pressure is abnormally high, resulting in problems such as a significant increase in cardiovascular risk. It is a serious, but an avoidable risk factor for coronary artery disease, hemorrhagic and ischemic stroke, and heart failure[1]. Long-term hypertension destroys blood vessels all over the body, especially in target organs like the heart, kidneys, brain, and eyes. Myocardial infarction, heart failure, renal failure, strokes, and vision impairment are the most common complications[2].

Blood pressure self-monitoring (BPSM) is the term for a patient’s regular use of a personal sphygmomanometer outside of a clinical context[3]. When a person takes their blood pressure outside of the clinic—at home, at work, or elsewhere—this is known as self-monitoring[4]. Self-monitoring of blood pressure was established in the 1930s and is currently used by about 10% of the general population in the United Kingdom[5].

Self-monitoring of blood pressure is an excellent technique to improve hypertension management and can be included to hypertensive patients’ routine care at regional hypertension management clinics[6].According to research, home blood pressure measures are more accurate than office blood pressure tests in terms of prediction accuracy[7]. When self-monitoring blood pressure, patients engage in self-care measures. Treatment adherence has improved and blood pressure has decreased as a result of these incidents. In hypertensive patients, BPSM is projected to become a standard aspect of their treatment[8].

Japanese hypertension guideline has explained that BPSM has several of benefits; highly reproducible, greater prognostic value, extremely effective for the evaluation of drug effects and their duration, used for telemedicine, facilitates long-term BP control, improves adherence to medications, detects seasonal variations and long-term changes in BP, essential for the diagnosis of white-coat hypertension and masked hypertension, detects morning and nighttime hypertension, important for the diagnosis and treatment of hypertension principally (in diabetes mellitus, pregnancy, children, and renal diseases), and has a great effect on the medical economy[9].

Blood pressure self-monitoring is becoming a fundamental part of hypertension management and primary care patients who self-initiated BPSM reported being more self-efficacious, but a lack of participation and guidance from their doctors generated confusion and hindered the true advantage of BPSM[10]. However, the act of discussing their BPSM readings with their health care providers gives rise to a greater doctor-patient therapeutic relationship[11].

Blood pressure self-monitoring could be an effective method to improve hypertension control and it could be integrated into the usual care of hypertensive patients in the hypertension management center of the community[6]. The practice of BPSM has numerous benefits to control of BP, improving the adherence rates to antihypertensive medications and approval of a better lifestyle[12].

The failure to comply with HTN’s self-care practice is the main reason for the poor control. Self-care activities have proven to be a significant and cost-effective intervention in the management and prevention of hypertension and its complications. Adherence to medicine, a low-fat diet, daily exercise, alcohol restriction, smoking cessation, weight loss, self-monitoring of blood pressure (BP), regular health checks, and stress reduction are all examples of hypertension self-care[13].

Several researchers have found that self-monitoring of blood pressure in hypertension patients varies between 24 and 82 percent in different European nations [14-17].Higher education, governmental employment, having an income of >3500 Ethiopian, duration of hypertension >6 years, having health insurance, having co-morbidities, receiving a health professional recommendation toward self-monitoring of BP, and having knowledge of hypertension-related complications were all found to be factors that were significantly associated with self-monitoring of BP in an Ethiopian study[18-23].

Because there is limited information from Ethiopia focusing on the study area, the study will fill the gap for blood pressure self-monitoring practice and associated factors among adult hypertensive patients in follow-up South Wollo Zone public hospitals.

## Methods and Materials

### Study area and period

The study was conducted from June 1 to June 30, 2022, at selected south Wollo zone public hospitals. There are a total of 14 public hospitals in the South Wollo zone serving about 4 million people, of which 11 of them are primary hospitals, 2 of them are general hospitals, and one is acomprehensive specialized hospital. The capital of the South Wollo zone is Dessie city, which is located 401 km far from Addis Ababa in the north-east of Ethiopia and has a multitude of health facilities, including two government hospitals, eight health centers, and three private general hospitals, two surgery centers, and ten higher clinics.

### Study Design

A facility-based cross-sectional study was conducted.

### Population

All hypertensive adults who visit public hospitals in South Wollo Zone were the source population, and all adult hypertensive patients on follow-up in the selected public hospitals during the data collection period were the study population.

### Inclusion and Exclusion criteria

All hypertensive patients who are above 18 years of age and taking antihypertensive drugs greater than or equal to 6 monthsand who have been on follow-up were included in the study. In contrast, patients who were unable to communicate and were severely ill were excluded from the study since they cannot provide valid information.

### Sample size determination

The sample size was determined using a single population proportion formula based on the following assumptions: considering a 95% of confidence level,3% margin of error, and 7.75% population proportion. research was done at Arsi zone blood pressure monitoring practice[1].

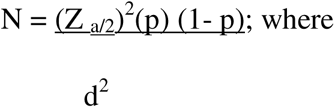

N: Sample size,

Z _a/2_ =1.96 (standardized normal distribution curve value for the 95% confidence Interval),

P = 0.0775 (proportion of good practice)

D = 0.05 (degree of margin of error)

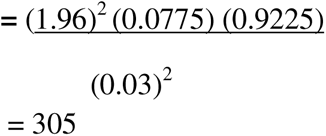

For the second objective, the sample size was calculated by using epi info version 7.2.5 stat Calc.Since the sample size calculated for the first objective was larger than the sample size calculated for the second objective, by adding a 10% non-response rate to the calculated sample size, the final sample will be 336.

### Sampling technique and procedures

From the total of 14 public hospitals located in the South Wollo Zone, 5 hospitals were selected randomly and the selected hospitals are Dessie comprehensive specialized hospital, Borumeda general hospital, Haik Primary hospital, Kelela Primary hospital, and Woreilu Primary hospital.

The study participants were chosen from each selected hospital using a systematic random sampling approach with a skip interval of 2 and the first study subject among the two was selected by lottery method. For each hospital, the proportionate allocation algorithm was used to determine the number of participants.

Allocating sampling proportional to the total population of each stratum using the formula:

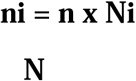

Where n=total sample size to be selected(336),

N=total population of all selected hospitals (840)

Ni= total population in each selected hospital

ni =sample size for each stratum

### Variables of the Study

#### Independent variables

##### Patient-related factors (socio-demographic, behavioral, and clinical factors)

- Age
- Sex
- Level of Education
- Occupation
- Family history of hypertension
- Duration of the disease
- Health insurance
- Co-morbidities
- History of HTN complication
- knowledge on hypertension self-care and complication
- Regular healthcare professional visits
- Smoking

##### Health care professional related factors

- Recommendations for using blood pressure self-monitoring
- Advice on the procedure of blood pressure self-monitoring
- Advice on type of device used for blood pressure self-monitoring

#### Dependent variable

- Practice of Blood pressure self-monitoring (good Vs poor)

### Operational definitions

#### A self-monitoring gadget

is a tool used to measure blood pressure on one’s own, without the aid of a medical professional[20].

#### Blood pressure Self-monitoring

is the self-measurement of blood pressure by patients at home using a self-monitoring device[20]. It was assessed by asking the question ‘Do you currently self-monitor your blood pressure (i.e. check your blood pressure by yourself using a self-monitoring blood pressure device at home)? [20]

#### Knowledge of hypertension self-care and complications

was assessed by asking nine knowledge questions with yes/no responses. By computing a mean score from answering correctly with yes = 1 and no = 0 ranging from 0 to 9 with a mean score of 5. Respondents were labeled to have[14, 24].

#### Good knowledge

if the study participants score 6 and above[14].

#### Poor knowledge

if the study participants score 6 and above (Error! Reference source not found.).

### Data collection tooland procedure

A semi-structured questionnaire wasprepared and utilized after studying relevant literature on the subject under investigation. For data collection, an Amharic language version of the questionnaire was employed. The data were collected by one health professional (clinical nurse) at each selected public hospital and one BSc nurse was assigned as a supervisor for each hospital. Data were collected through face-to-face interviews by the data collector; supervisors check on the spot to ensure its completeness. The patient was initially asked about his or her interest in taking part in the study. Once a qualified data collector has finished the service, the patient who consents to participate was subsequently questioned.

### Data quality control

To ensure uniformity, the questionnaire was translated from English to Amharic by a language expert translator and then back to English by a second expert translator who is a health professional. The questionnaire was pretested on 5% (17) of the overall sample size of hypertension patients on follow-up at Kombolcha primary hospital, with any necessary revisions made before it is used for actual data collection. The study equipment and data collection process were taught to data collectors and supervisors over the course of two days. The collected data werechecked for completeness by the primary investigator and supervisors.

### Data processing and analysis

The information obtained was double-checked, coded, and entered into Epi-Data 4.6. The data were then exported to version 25 of the Statistical Package for the Social Sciences (SPSS) for statistical analysis.The study participants were described using descriptive statistics such as frequency, percentage, and measures of central tendency. Tables and text were used to present the data. Then, to identify factors associated with BP self-monitoring, a binary logistic regression model was used. To examine the relationship between dependent and independent factors, all independent variables with a p-value of ≤0.25were included in the multivariable logistic regression model. Using multivariable logistic regression analysis, a P-value of 0.05 was used to declare as statisticallysignificant; an AOR with a 95% confidence interval was used to identify factors significantly associated with BP self-monitoring.Model fitness was checked by Hosmer and Lemshow statistic and the P-value obtained was 0.92 and multicollinearity was checked by using variance inflation factor (VIF).

## Results

### Socio-demographic characteristics of respondents

A total of 336 respondents were enrolled in the study making a response rate of 100%. The mean age of the participants was 43 years with SD of ±14 years. More than half 196 (58.3%) and 199 (59.2%) of the respondents were males and urban dwellers respectively. The majority 235(69.9%) of the respondents were married. Regarding educational status, 178(53.0%) of the respondents have attended above secondary education. Regarding average monthly income, nearly one-third 123(36.6%) of the respondents had ≥ 3000 Ethiopian birr average monthly income (Table 1).

**Table 1:** Socio-demographic characteristics of hypertensive patients who were attending public hospitals in South Wollo Zone, Northeast Ethiopia, 2022 [n=336]

| Variable | Response | Frequency | Percent |
| --- | --- | --- | --- |
| Sex of respondents | Male | 196 | 58.3 |
|  | Female | 140 | 41.7 |
| Religion of respondents | Orthodox | 150 | 44.6 |
|  | Muslim | 161 | 47.9 |
|  | Protestant | 19 | 5.7 |
|  | Others | 6 | 1.8 |
| Educational status of respondents | Illiterate | 29 | 8.6 |
|  | primary education | 18 | 5.4 |
|  | secondary education | 111 | 33.0 |
|  | above secondary education | 178 | 53.0 |
| Marital status of respondents | Single | 63 | 18.8 |
|  | Married | 235 | 69.9 |
|  | Divorced | 26 | 7.7 |
|  | Widowed | 12 | 3.6 |
| Residence | Urban | 199 | 59.2 |
|  | Rural | 137 | 40.8 |
| Average monthly income | <1000 ETB | 54 | 16.1 |
|  | 1000-1999 ETB | 59 | 17.6 |
|  | 2000-2999 ETB | 100 | 29.8 |
|  | >=3000 ETB | 123 | 36.6 |

### Behavioral and health related characteristics of participants

Among the participants, nearly one third 130 (38.7%) had health insurance. The duration of their disease for 238 (70.8%) respondents was five years or less since its diagnosis by health professionals. Regarding behavior, 77(22.9%) of them were currently smoking and 246 (73.2%) of the respondents had regular follow-upaccording to their appointment (Table 2).

**Table 2:**
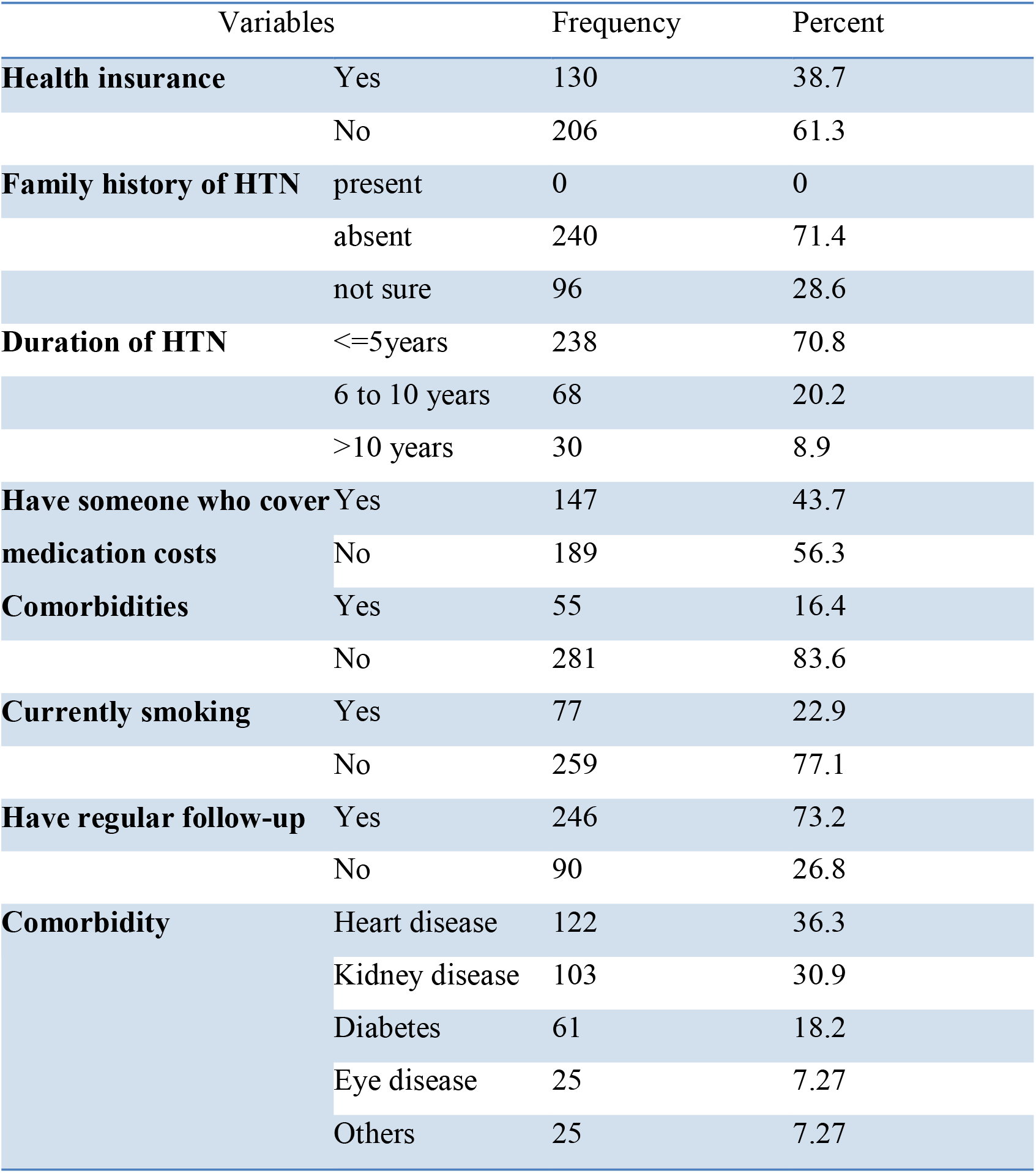
Behavioural and health-related characteristics of hypertensive patients who were attending public hospitals in South Wollo Zone, Northeast Ethiopia, 2022 [n=336]

| Variables |  | Frequency | Percent |
| --- | --- | --- | --- |
| **Health insurance** | Yes | 130 | 38.7 |
|  | No | 206 | 61.3 |
| **Family history of HTN** | present | 0 | 0 |
|  | absent | 240 | 71.4 |
|  | not sure | 96 | 28.6 |
| **Duration of HTN** | <=5years | 238 | 70.8 |
|  | 6 to 10 years | 68 | 20.2 |
|  | >10 years | 30 | 8.9 |
| **Have someone who cover medication costs** | Yes | 147 | 43.7 |
|  | No | 189 | 56.3 |
| **Comorbidities** | Yes | 55 | 16.4 |
|  | No | 281 | 83.6 |
| **Currently smoking** | Yes | 77 | 22.9 |
|  | No | 259 | 77.1 |
| **Have regular follow-up** | Yes | 246 | 73.2 |
|  | No | 90 | 26.8 |
| **Comorbidity** | Heart disease | 122 | 36.3 |
|  | Kidney disease | 103 | 30.9 |
|  | Diabetes | 61 | 18.2 |
|  | Eye disease | 25 | 7.27 |
|  | Others | 25 | 7.27 |

### Health care professional related factors

Of the total respondents, nearly half 166(49.4%) of the respondents had been recommended by health professionals toward using BP self-monitoring; around one-third 116(34.5%) were advised on the procedure of BP self-monitoring, and 256(76.2%) of them were told about hypertension-related target organ complications by a health care professional during follow-up (Table 3).

**Table 3:**
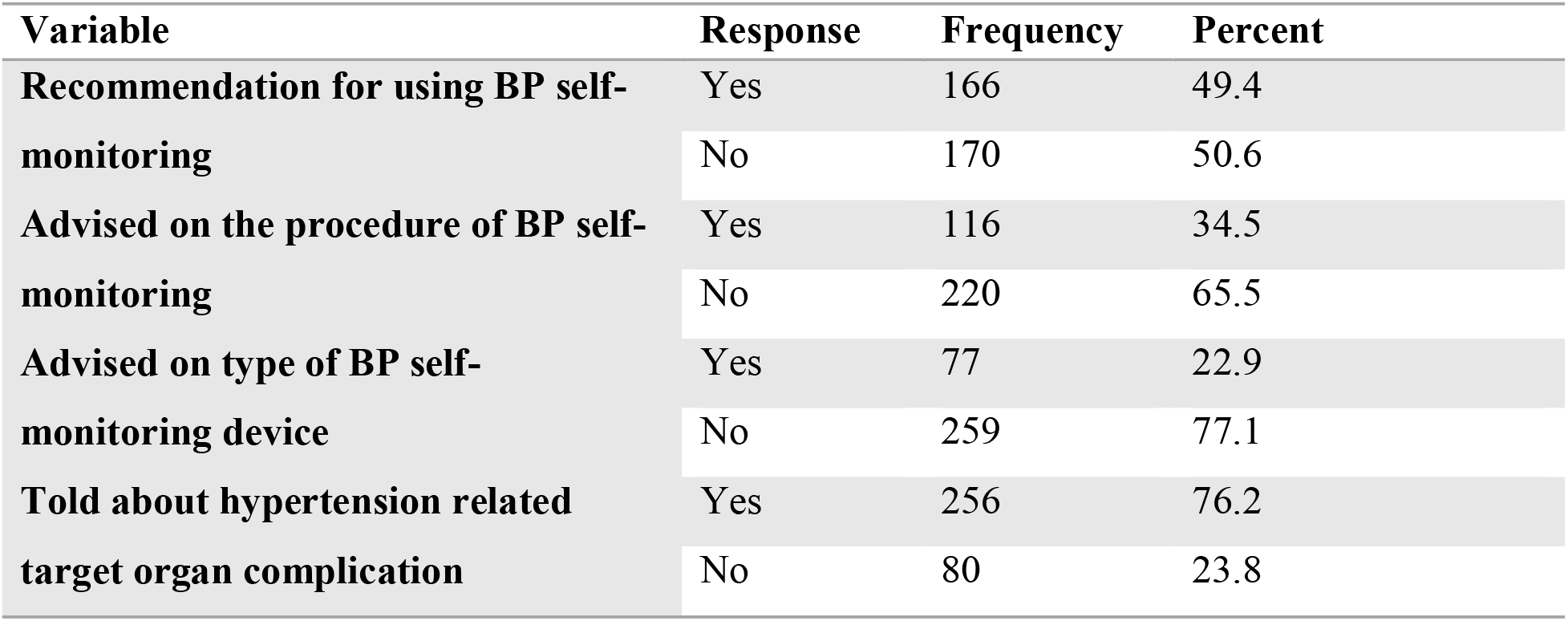
Health care professional related factors among hypertensive patients who were attending public hospitals in South Wollo Zone, Northeast Ethiopia, 2022 [n=336]

| Variable | Response | Frequency | Percent |
| --- | --- | --- | --- |
| **Recommendation for using BP self-monitoring** | Yes | 166 | 49.4 |
|  | No | 170 | 50.6 |
| **Advised on the procedure of BP self-monitoring** | Yes | 116 | 34.5 |
|  | No | 220 | 65.5 |
| **Advised on type of BP self-monitoring device** | Yes | 77 | 22.9 |
|  | No | 259 | 77.1 |
| **Told about hypertension related target organ complication** | Yes | 256 | 76.2 |
|  | No | 80 | 23.8 |

### Knowledge of hypertension self-care and complications

Among the total of 336 hypertensive patients, 85 (25.3%) had good knowledge of hypertension self-care and complications.

### Blood pressure self-monitoring practice

Three hundred thirty-six (336) hypertensive patients were invited to answer the question ‘Do you currently self-monitor your blood pressure (i.e. check your Blood Pressure by yourself using a self-monitoring blood pressure device at home)?’ and it was found that 31(8.93%) of them use personal BP device to monitor their BP at home.

### Factors associated with BP self-monitoring practice

In bi-variable logistic regression analysis, age, sex, residence, health insurance, comorbidity, regular healthcare professional visit, recommendation toward BP self-monitoring, advice on the procedure of BP self-monitoring, advice on the type of devices used for BP self-monitoring, and knowledge on hypertension self-care were associated with BP self-monitoring practice at P≤0.25. However, only residence, comorbidity, regular healthcare professional visits, advice on the type of devices used for BP self-monitoring, and knowledge on hypertension self-care were significantly associated with BP self-monitoring practice at P < 0.05.

Keeping other variables in the model constant, the odds of BP self-monitoring among participants who have comorbidities were nearly five times [AOR=4.80, 95%CI (1.23, 18.69)] higher than participants who had no comorbidity. Patients living in urban areas were four times [AOR=3.97, 95%CI (1.11, 14.20)] more likely to have blood pressure self-monitoring practice as compared to patients living in rural areas. Similarly, patients having regular follow-upwere nearly five times [AOR=4.64, 95%CI (1.02, 21.14)] more likely to have BP self-monitoring practice as compared to patients having no regular follow-up. Keeping other variables in the model constant, being advised by health professionals on the type of device used for BP self-monitoring increases the odds of BP self-monitoring by fivefold [AOR=5.26, 95%CI (1.49, 18.58)] compared to patients not advised by health professionals on type of device used for BP self-monitoring. Moreover, patients having good knowledge about hypertension self-care and complicationswere thirteen times more likely to have blood pressure self-monitoring practice as compared to patients having poor knowledge about hypertension self-care and complications. [AOR=13.13, 95%CI (4.21, 40.99)](Table 4).

**Table 4:** Factors associated with BP self-monitoring practice among hypertensive patients who were attending public hospitals in South Wollo Zone, Northeast Ethiopia, 2022 [n=336]

| Variables | Category | BP self-monitoring practice |  | COR (95%CI) | AOR (95%CI) |
| --- | --- | --- | --- | --- | --- |
|  |  | Yes | No |  |  |
| Age of respondent |  |  |  | 0.98(0.95,1.00) | 0.97(0.94,1.01) |
| Sex of respondent | Male | 14 | 182 | 0.59(0.28,1.26) | 0.84(0.29,2.40) |
|  | Female | 16 | 124 | 1 | 1 |
| Residence | Urban | 25 | 174 | 3.79(1.41,10.17) | 3.97(1.11,14.20) |
|  | Rural | 5 | 132 | 1 | 1 |
| Health insurance | Yes | 8 | 122 | 0.55(0.24,1.27) | 2.05(0.14,28.93) |
|  | No | 22 | 184 | 1 | 1 |
| Someone who covers your medicine | Yes | 9 | 138 | 0.52(0.23,1.18) | 0.40(0.03,5.22) |
|  | No | 21 | 168 | 1 | 1 |
| Comorbidities | Present | 8 | 47 | 2.00(0.84,4.77) | 4.80(1.23,18.69) |
|  | Absent | 22 | 259 | 1 | 1 |
| Regular follow-up | Yes | 25 | 221 | 1.92(0.71,5.18) | 4.64(1.02,21.14) |
|  | No | 5 | 85 | 1 | 1 |
| Recommendation on BP self-monitoring | Yes | 25 | 141 | 5.85(2.18,15.68) | 0.65(0.11,3.97) |
|  | No | 5 | 165 | 1 | 1 |
| Advice on the procedures of BP self-monitoring | Yes | 25 | 91 | 11.81(4.38,31.82) | 5.12(0.81,32.40) |
|  | No | 5 | 215 | 1 | 1 |
| Advice on type of device used for BP self-monitoring | Yes | 22 | 55 | 12.55(5.31,29.66) | 5.26(1.49,18.58) |
|  | No | 8 | 251 | 1 | 1 |
| Knowledge about hypertension self-care | Good | 25 | 60 | 20.50(7.54,55.77) | 13.13(4.21,40.99) |
|  | Poor | 5 | 246 | 1 | 1 |

## Discussion

The proportion of BP self-monitoring among hypertensive patients in this study was 8.93% [95%CI; 6.3, 12.5]. This finding is lower when compared to studies conducted in America, Czech Republic, United States, Canada, and Italy the reported proportions of 53.8%, 40%, 41.6%, 50%, and 74.7% respectively[21, 22, 25-27]. This variation might be due to differences in sample size, socio-economic status of population, and sampling procedure since the study in America was undertaken with an online survey, patients in low-income category might not be involved due to service inaccessibility which could overestimate the proportion of BP self-monitoring. The other reason for the low proportion of BP self-monitoring obtained in this study as compared to a study conducted in Czech Republic is the population segment difference where the study was conducted among 552 hypertensive patients aged (25-75 years). Study design and study setting differences for United States study where a cross-sectional, correlational design was used among urban community population, study setting differences for Canada study where the study conducted among community pharmacies hypertensive patients and differences in sample size for study in Italy where it was conducted among 855 hypertensive patients.

This result is also lower than findings reported from studies done in west Midlands (UK), Muscat (governorate of Sultanate of Oman), and Amman (Jordan) that shows the proportion of BP self-monitoring; 30.7%, 40%, and 82% respectively[12, 23, 26]. The variation might be due to differences in sample size (1815 for the study done at west Midlands, UK), the difference in study setting (a study in Jordan was conducted among institutions in Amman, the capital city of Jordan, and the pharmacist participation in counseling patients on the proper use of blood pressure monitors and delivering needed relevant education in addition to other health care professional as a study report)

This finding was also lower than results reported from a study done in Karachi (southern Asia), northeastern Singapore (Asia), northern Carolina, and China where the prevalence of BP Self-Monitoring among hypertensive patients was 25%, 24%, 43.1% and 24.5% respectively[12, 26, 28, 29]. The variation might be due to differences in study setting where the study was conducted at the tertiary hospital for the study of Karachi, Southern Asia and differences in sample size (700) for the study done at Northern Carolina.

Adviceon type of devices used for BP self-monitoring was an important significant factor for BP self-monitoring.Respondents who were advisedonthe type of devices used for BP self-monitoring were five times more likely to have BP self-monitoring practice as compared to those who were not advised. This finding was supported by a study done inUnited States, Northern Carolina, and Arsi Zone (Ethiopia)[12, 19, 21]. The reason might be since having awareness about BP self-monitoring devices can increase the demand of blood pressure self-monitoring which finally improves BP self-monitoring practice.

Patients who had good knowledge about hypertension self-care and related target organ complications were thirteen times more likely to have BP self-monitoring when compared to their contraries. This result was consistent with studies conducted in the United States[28] and Arsi Zone Southeastern Ethiopia[19]. The reason might be due to the fact that having knowledge of hypertension-related organ complications will make the patient more conscious about the seriousness of the disease and the patient might be tensioned to control the disease and focuses on strategies of managing the disease like BP self-monitoring because knowing complications make the patient think that he/she will die due to the complications of the disease if they are not controlled and knowing about hypertension self-care will also create confidence and self-efficacy for using self-monitoring devices.

Those patients who had co-morbidities were nearly five times more likely to have BP self-monitoring practice than those having co-morbidities. This result was supported by a study done at Arsi Zone Southeastern Ethiopia[19]. The reason might be due to the fact that the presence of comorbidity increased concern about controlling the disease which intern leads to improved health facility visits, increased knowledge of hypertension self-care and increased chance to get health professionals counseling these finally leads to increased blood pressure self-monitoring practice.

Those patients who had regular follow-up (regular health facility visit) were nearly five times more likely to have BP self-monitoring practice than those having no regular health follow-up. This result was supported by an online survey report conducted in United States of America[14]. The reason might be due to the fact that having regular follow-up is associated with increased knowledge of hypertension self-care which can increase the practice of Blood Pressure self-monitoring.

Urban residents were nearly four times more likely to have BP self-monitoring than those who were rural residents. The possible reason is that living in an urban area increased information access which inturn increased knowledge of hypertension self-care which can increase the practice of Blood Pressure self-monitoring.

## Conclusion

The proportion of BP self-monitoring among hypertensive patients who were attending public hospitals in South Wollo Zone, Northeast Ethiopia was low. Having urban residence, comorbidity, regular healthcare professional visits, advice on the type of devices used for Blood Pressure self-monitoring, and knowledge of hypertension self-care were factors significantly associated with Blood Pressure self-monitoring practice.Health care providers should focus on hypertensive patients who had co-morbidities, no regular healthcare professional visits, and who had no adequate knowledge regarding hypertension self-care and complications. Awareness creation programs should be set and the patients should be taught on hypertension-related complications and BP self-monitoring practice procedures and the precautions required during utilization. Furthermore, they should inform the patient to self-monitor their BP. The Zone health department should encourage and design awareness creation programs regarding BP self-monitoring into hypertension self-care programs by using different mainstream Media which considers accessibility to the rural community. Further research with a strong study design and multisite is suggested for researchers.

## Data Availability

All data produced in the present study are available upon reasonable request to the authors

## Ethical considerations

Ethical clearance was obtained from the Ethical Review Committee of the Tropical College of Medicine Dessie Campus. Then, the supportive letter was obtained from all selected public hospitals to obtain permission and conduct the study. After explaining the objective of the study, oral informed consent was taken from each study participantbefore data collection. To maintain confidentiality, the patient name was not included in the questioner.

### Abbreviation and Acronyms

AOR: Adjusted Odds Ratio
BP: Blood Pressure
BPSM: Blood Pressure Self-Monitoring
CI: Confidence Interval
COR: Crude Odds Ratio
HBPM: Home Blood Pressure Monitoring
SBP: Systolic Blood Pressure
SPSS: Statistical Package for Social Science

## Consent for publication

“Not applicable”

## Availability of data and materials

The dataset will not be shared in order to protect the participants’ identitiesbut is available from the corresponding author onreasonable request.

## Author’s contribution

**EG** conceived and designed the study, performed analysis and interpretation of data. AE and ZM advised andsupervised the design conception, analysis, interpretation of data and made critical comments at each step of research. BT, SA and TL drafted the manuscript. All authors read and approved the final Manuscript. Confidentiality and anonymity were ensured throughout the execution of the study.

## Acknowledgement

We want to forward our heartfelt thanks to the study participants and data collectors for their commitment and cooperation during data collection period. We would also like to thank South Wollo Zone Health Department and its staffs for their support.

